# The Effect of Gender on Covid-19 Infections and Mortality in Germany: Insights From Age- and Sex-Specific Modelling of Contact Rates, Infections, and Deaths

**DOI:** 10.1101/2020.10.06.20207951

**Authors:** Achim Dörre, Gabriele Doblhammer

**Affiliations:** Department of Economics, University of Rostock, Germany; Department of Sociology and Demography, University of Rostock, Germany

## Abstract

**Background:** Recent research points towards age- and sex-specific transmission of COVID-19 infections and their outcomes. The effect of sex, however, has been overlooked in past modelling approaches of COVID-19 infections.

**Aim:** The aim of our study is to develop an age- and sex-specific model of COVID-19 transmission and to explore how contact changes effect COVID-19 infection and death rates.

**Method:** We consider a compartment model to establish forecasts of the COVID-19 epidemic, in which the compartments are subdivided into different age groups and genders. Estimated contact patterns, based on other studies, are incorporated to account for age- and sex-specific social behaviour. The model is fitted to real data and used for assessing hypothetical scenarios with regard to lockdown measures.

**Results:** Under current mitigation measures as of mid-August, active COVID-19 cases will double by the end of October 2020. Infection rates will be highest among the young and working ages, but will also rise among the old. Sex ratios reveal higher infection risks among women than men at working ages; the opposite holds true at old age. Death rates in all age groups are twice as high among men as women. Small changes in contact rates at working and young ages may have a considerable effect on infections and mortality at old age, with elderly men being always at higher risk of infection and mortality.

**Discussion:** Our results underline the high importance of the non-pharmaceutical mitigation measures in low-infection phases of the pandemic to prevent that an increase in contact rates leads to higher mortality among the elderly. Gender differences in contact rates, in addition to biological mechanisms related to the immune system, may contribute to sex-specific infection rates and their mortality outcome. To further explore possible pathways, more data on COVID-19 transmission is needed which includes socio-demographic information.

## 1 Introduction

Right from the start of the COVID-19 pandemic, the importance of age on COVID-19 contraction and fatality has been recognised (among others, Esteve et al. (2020), Dudel et al. (2020), Kulu and Dorey (2020), Wu and McGoogan (2020), Karagiannidis et al. (2020)), as well as of coresidence patterns (Esteve et al. (2020)). Compartment and agent-based models aiming at projecting the spread of the disease have incorporated age as an important variable of transmission (e.g. Davies et al. (2020), Deforche (2020), Colombo et al. (2020), Blyuss and Kyrychko (2020), Balabdaoui and Mohr (2020)), in addition to other characteristics such as space (Colombo et al. (2020)) or contact patterns (Zhang et al. (2020)). An important determinant, which appeared to be largely overlooked in modelling exercises, is sex. In the following, we will refer to sex when discussing technical details and biological factors, and gender, when referring to social factors. While studies generally notice that infection and in particular fatality rates were higher among elderly men than women, the reverse appears to be true for infections at working ages (Sobotka et al. (2020)). In Germany, during the first wave of the pandemic through mid-May, infection rates were higher among women than men at working ages (Figure 1), while they were higher for men thereafter.

**Figure 1:**
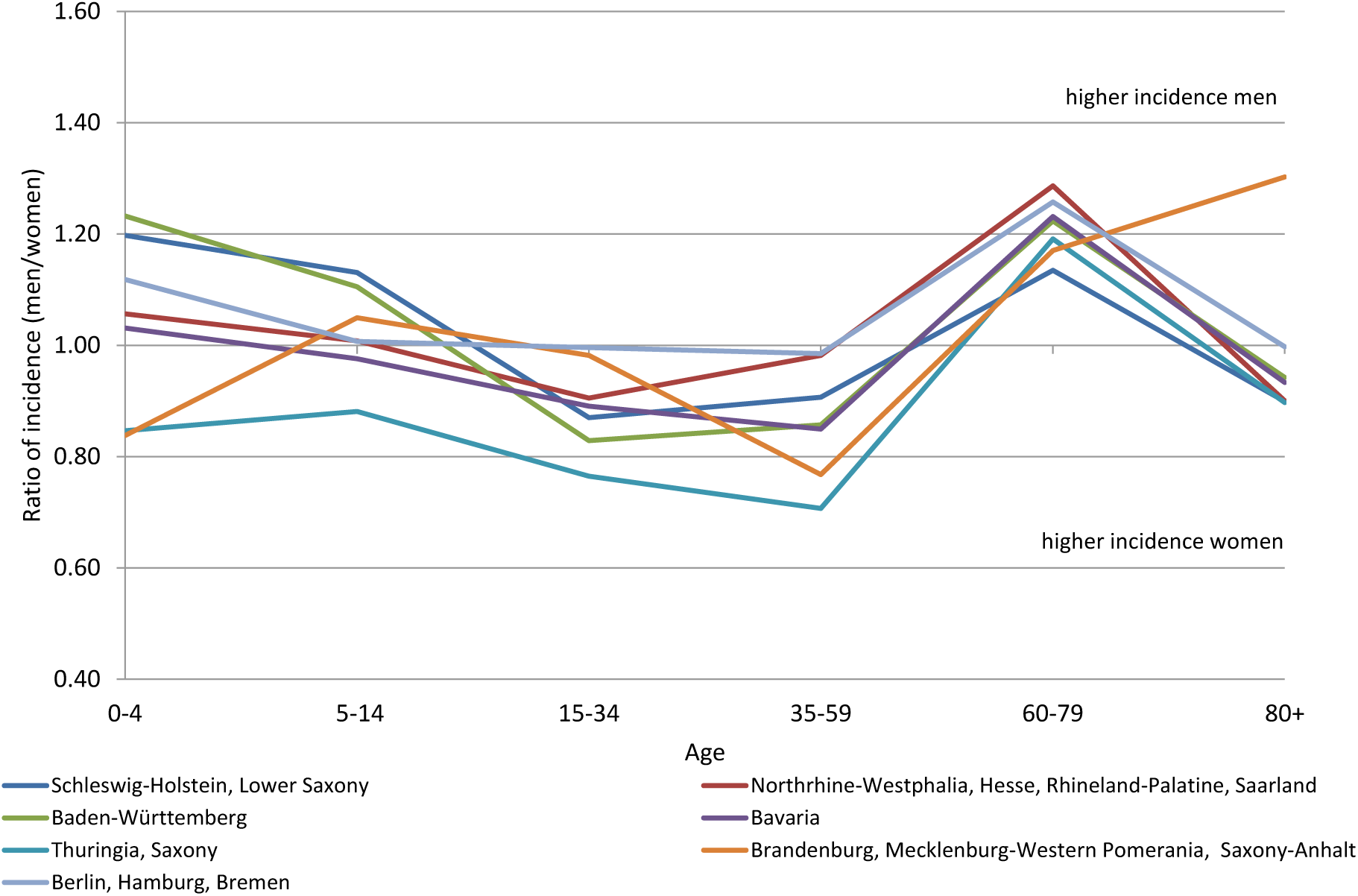
Sex ratio (male/female) of COVID-19 incidence through 15 May 2020 for German Laender by age, Data Source: Robert Koch-Institut Dashboard, authors’ calculations

One reason for this difference, in addition to biological factors (see discussion below) may lie in gender-specific contact rates. Estimates of contact rates (van de Kassteele et al. (2017)) based on the POLYMOD study (Mossong et al. (2008)) showed that household, workplace and school structures strongly shape age- and gender-specific contacts made by individuals. Using the contact matrices from the latter study and calculating the ratio of the age-specific number of contacts for men and women (contacts men/contacts women) a clear pattern emerges (Figure 2): among ages 20–39, contacts are between 13%–26% higher among women, while among ages 50 to 69, they are 9%–14% higher among men. At the highest ages, the pattern reverses again, with women having slightly more contacts.

**Figure 2:**
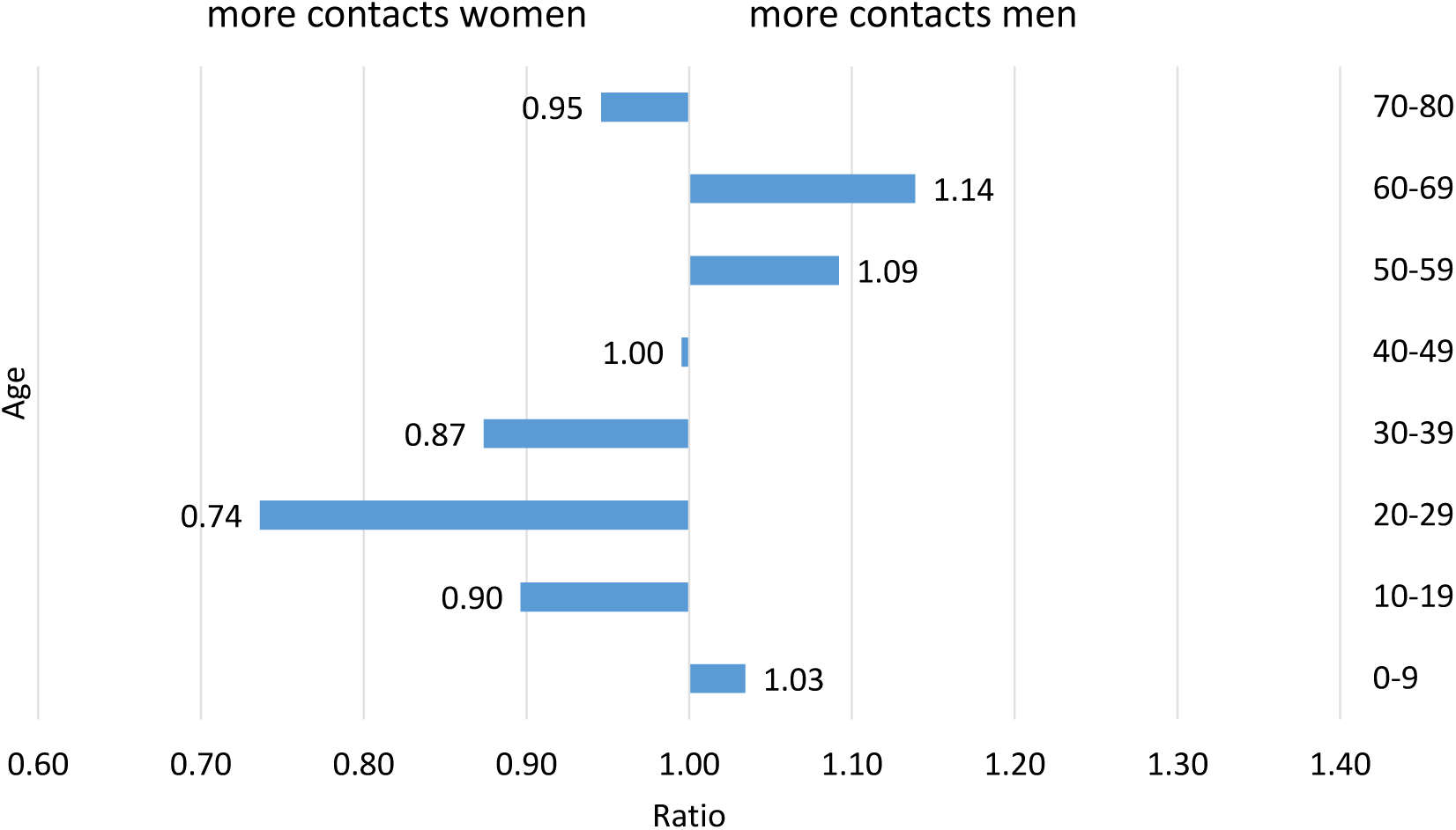
Ratio of the average number of contacts among men compared to women, Data Source: (van de Kassteele et al. (2017)).

The aim of our study is to model COVID-19 transmission taking into account the two crucial demographic factors age and sex. We develop an SEIRD-model that incorporates age- and sex-specific contacts, which shape transmission rates. The model may be used for short- and long-term projections, our example explores short-term effects up to two and a half months of hypothetical changes in contact rates. The model can be used to develop scenarios which address the effects of age- and gender-specific changes in contacts due to the closing of schools, kindergarten and shops, or work in home office, as well as to explore the effect lifting of these measures. While we are not able to address these effects separately, we translate them into hypothetical changes in age- and sex-specific contact rates by developing four scenarios. The first scenario reflects a continuation of the situation of mid-August 2020; the second assumes a lifting of measures mainly at working ages, and the third extends this to children, adolescents, and young adults. The manuscript is structured as follows: First we introduce the basic SEIRD model and discuss how age- and sex-specific contact modelling was incorporated. We present the numerical implementation of the model, model fitting and the development of uncertainty intervals. Then we introduce our scenarios and present the projection results in terms of number of active infections (prevalence), and cumulated number of deaths by 31 October 2020. We also explore how increasing contacts affect sex-ratios in infections and deaths. We close with a discussion of the results, the strengths and limitations of our model, as well as policy implications.

## 2 Methods

The core of the epidemiological model is an SEIRD compartment model (see Hethcote (2000)) consisting of the epidemiological states *S* (susceptible, i.e. not yet exposed to the virus), *E* (exposed, but not infectious), *I* (infectious), *R* (recovered), and *D* (dead). The compartments represent individual states with respect to contagious diseases, i.e. COVID-19 in this case, and the transitions between them are considered on a population level (see Figure 3). In this sense, the compartment model is used to describe a population process, but is not intended to model individual processes with respect to COVID-19.

**Figure 3:**
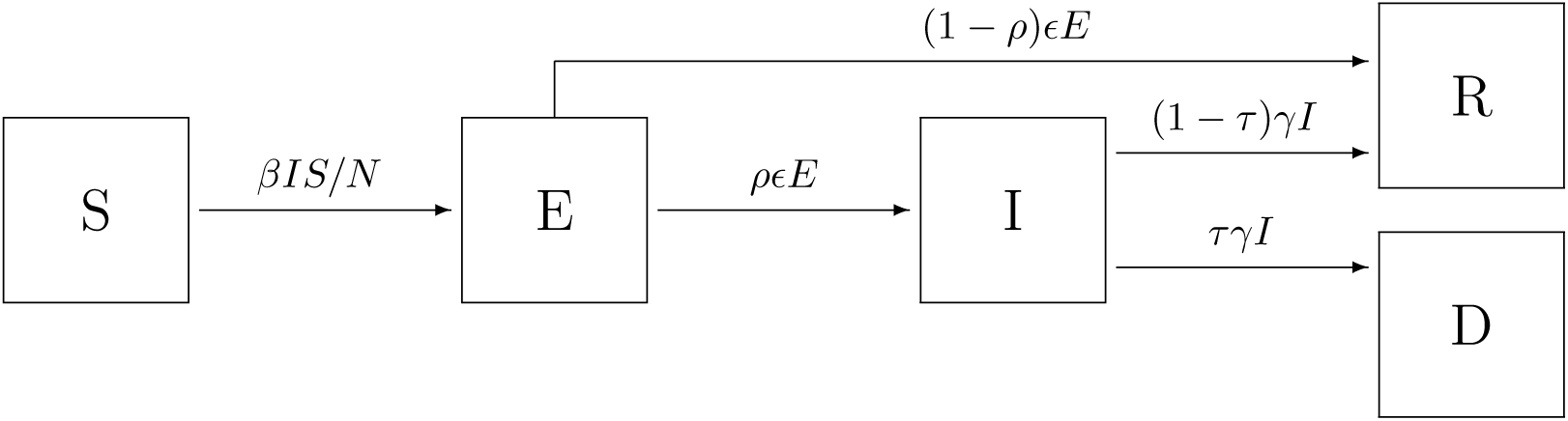
SEIRD compartment model with 5 transitions. (*S* → *E*: susceptible person becomes exposed to the virus, *E* → *I*: exposed person becomes infectious, *E* → *R*: exposed person is removed due to recovery, *I* → *R*: infectious person is removed due to recovery, *I* → *D*: infectious person is removed due to death)

The following essential rate and fraction parameters are involved in the model:

1. *β* (contact rate): the average number of individual contacts per specified timespan that are potentially sufficient to transmit the virus (see below for detailed specification)
2. *ρ* (manifestation index, fraction): the fraction of people who become infectious at some time after being exposed to the virus
3. *ϵ* (incubation rate): the mean rate of exposed people to become infectious; 1*/∈* is the average incubation time
4. *γ* (recovery rate): the mean rate of exiting the infectious state, either to recovery or death; 1*/γ* is the average duration of the disease
5. *τ* (infection fatality rate): the fraction of people who die due to COVID-19

### 2.1 Contact modeling

The contact model is considered for a population of *N* individuals, which is decomposed into *A* disjoint groups. For each group *a* = 1, …, *A*, the proportion of individuals with regard to the whole population is *N*_*a*_*/N*, where *N*_*a*_ denotes the number of individuals in group *a*. For any *a* ∈ {1, .., *A*} and *b* ∈ {1, …, *A*}, let *λ*_*ab*_ be the average number of contacts of an arbitrary individual from group *a* with individuals in group *b* during a fixed base time unit *δ*, e.g. 24 hours.

More specifically, define *η*_*ab*_(*t*_1_, *t*_2_) as the random number of contacts of an individual in group *a* with any individual from group *b* over the timespan [*t*_1_, *t*_2_] and 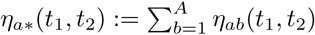 as the (random) overall number of contacts of an individual from group *a*. It is assumed that *η*_*ab*_(*t*_1_, *t*_2_) is Poisson distributed as

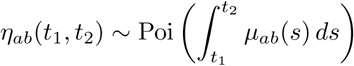

via the contact intensity *µ*_*ab*_(*t*). By assuming independence of contacts to different groups, it follows that *η*_*a∗*_(*t*_1_, *t*_2_) is also Poisson distributed having intensity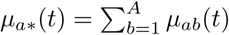. The average rate of contact of any individual from group *a* with group *b* is then obtained as

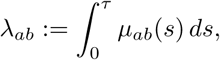

where for the sake of simplicity we assume that *µ*_*ab*_(*t*) is periodic in the sense that *µ*_*ab*_(*t* + *δ*) = *µ*_*ab*_(*t*) for all *t ≥* 0. Deviations from these assumptions can be incorporated by appropriate modifications to the contact model and parameter set. In the compartment modeling approach, individuals within each group are generally assumed to be homogenous with respect to contact behaviour and no individual effects are considered.

### 2.2 Group-specific system of ODEs

In order to address the potential impact of the implementation and easing of lockdown measures, we expand the model structure to group-specific compartments. Below, we define groups according to sex and age group, but the following reasoning is valid for any specification of disjoint groups, given that the resulting groups are sufficiently large. Specifically, for given groups *a* = 1, …, *A* and any time *t*, set *S*_*a*_(*t*) as the number of susceptible people in group *a* at time *t, E*_*a*_(*t*) as the number of exposed people in group *a* at time *t*, and so on. The group-specific compartment model is characterised by the ODE system

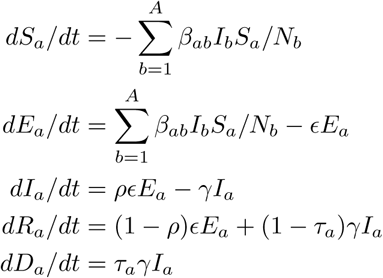

for all groups *a* = 1, …, *A*, which is a direct extension of the ODE system of the basic compartment model for the special case *A* = 1. We define

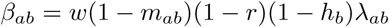

as the effective contact rate between groups *a* and *b*, where *w* is the secondary attack rate, *m*_*ab*_ is the specific mitigation effect by lockdown measures with regard to contacts between groups *a* and *b, r* is a general factor that accounts for compliance to distance, isolation and quarantine orders, *h*_*b*_ is the proportion of infectious people in group *b* in need of hospitalisation and *λ*_*ab*_ is the basic contact rate between groups *a* and *b* when no lockdown measures are in place. As we are primarily interested in short-term prediction, we do not model biological aging, i.e. transitions between demographic groups. Therefore, f or any time *t*, com partment-specific add itivity is assumed, i.e. *S*(*t*) = ∑_*a*_ *S*_*a*_(*t*), *E*(*t*) = ∑_*a*_ *E*_*a*_(*t*), *I*(*t*) = ∑_*a*_ *I*_*a*_(*t*), *R*(*t*) = ∑_*a*_ *R*_*a*_(*t*) and *D*(*t*) = ∑_*a*_ *D*_*a*_(*t*) and *N* = *S*(*t*) + *E*(*t*) + *I*(*t*) + *R*(*t*) + *D*(*t*). The system is closed, meaning that the sum of all ODEs is 0 at each time *t*.

In the absence of any lockdown measures, the general contact patterns are characterised by the basic contact rates *λ*_*ab*_, which represent how intensive/often group *a* has any contact with group *b* sufficient for potential virus transmission. In the POLYMOD study (Mossong et al. (2008), 7,290 participants from 8 countries including Germany reported the number and extent of their social contacts during a randomly assigned 24 hour period, using a written diary. The age and gender of the contacted persons were recorded, among other information. Overall, the study contains information on 97,904 contacts, distributed across the 8 participating countries. The overall contact pattern for Germany is displayed in Figure 4.

**Figure 4:**
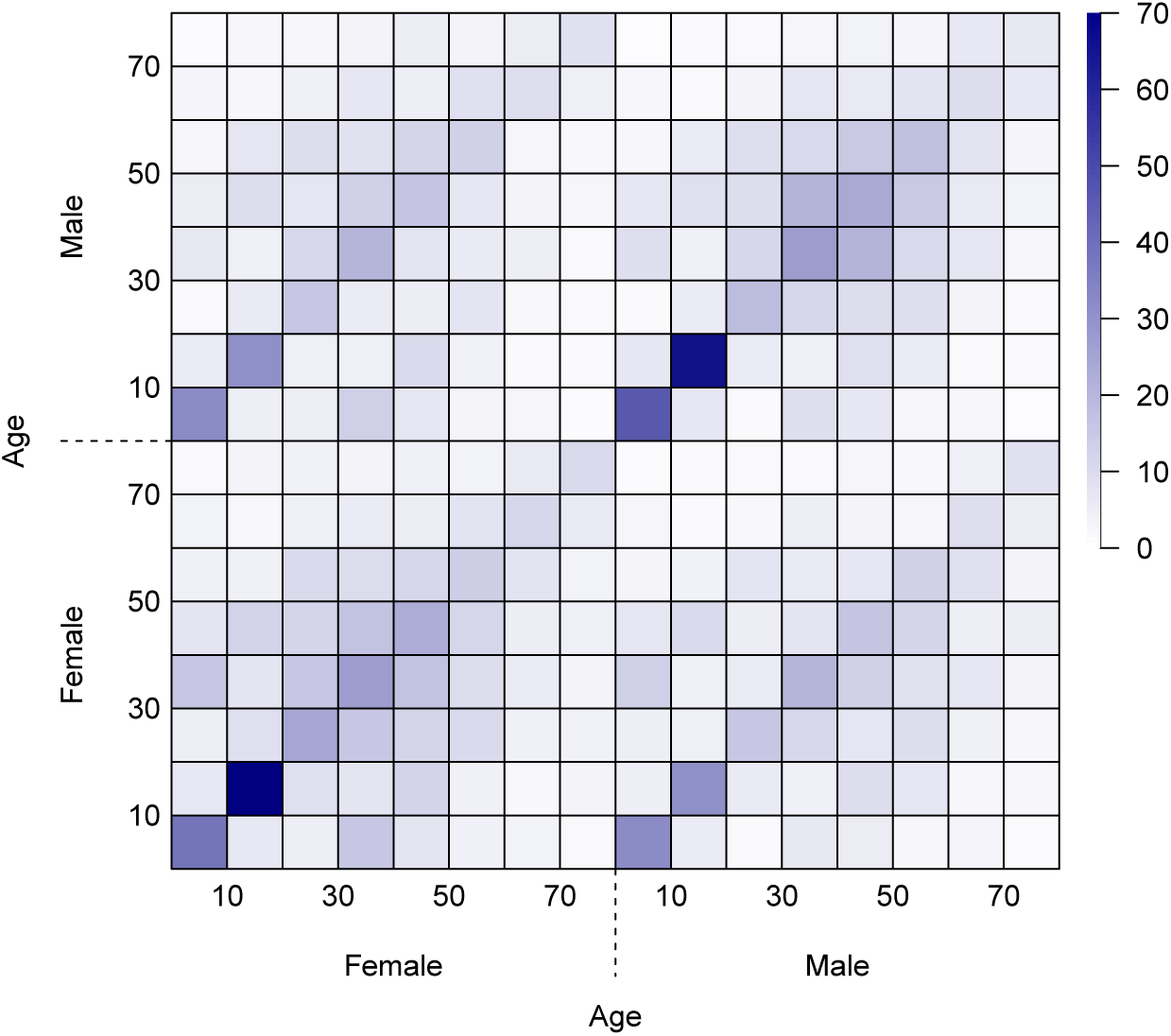
Overall contact rates *λ*_*ab*_ in Germany for different sex and age groups in the absence of lockdown measures (based on van de Kassteele et al. (2017); the scale displays the average number of contacts over the course of 24 hours).

**Figure 5:**
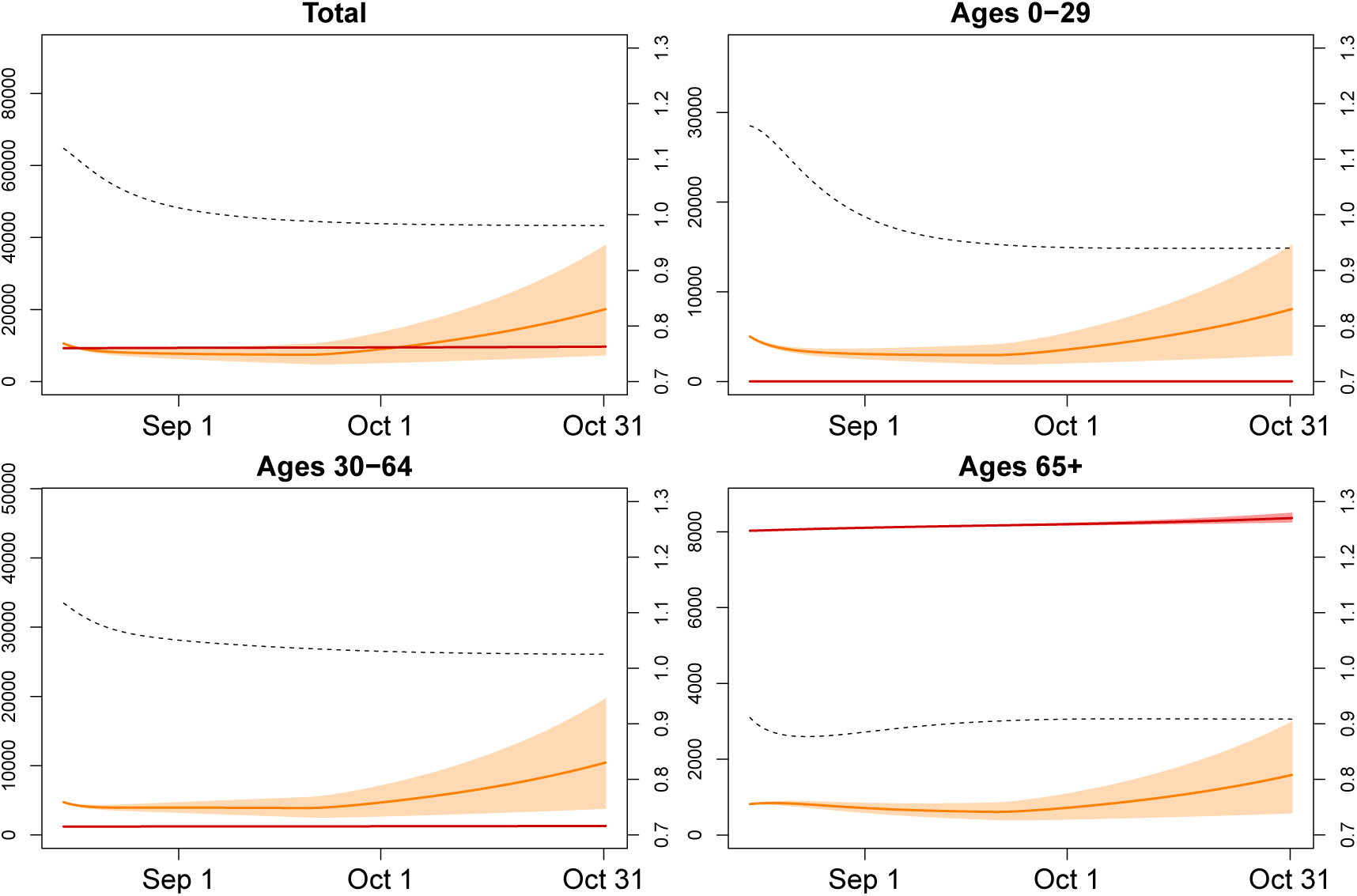
Scenario 1: Ongoing lockdown measures comparable to mid-August (solid orange line: mean number of active infections at time t, solid red line: mean cumulative number of deaths until time *t*, intervals represent 80% range due to parameter uncertainty, dashed line: male/female ratio of infections at time *t*).

**Figure 6:**
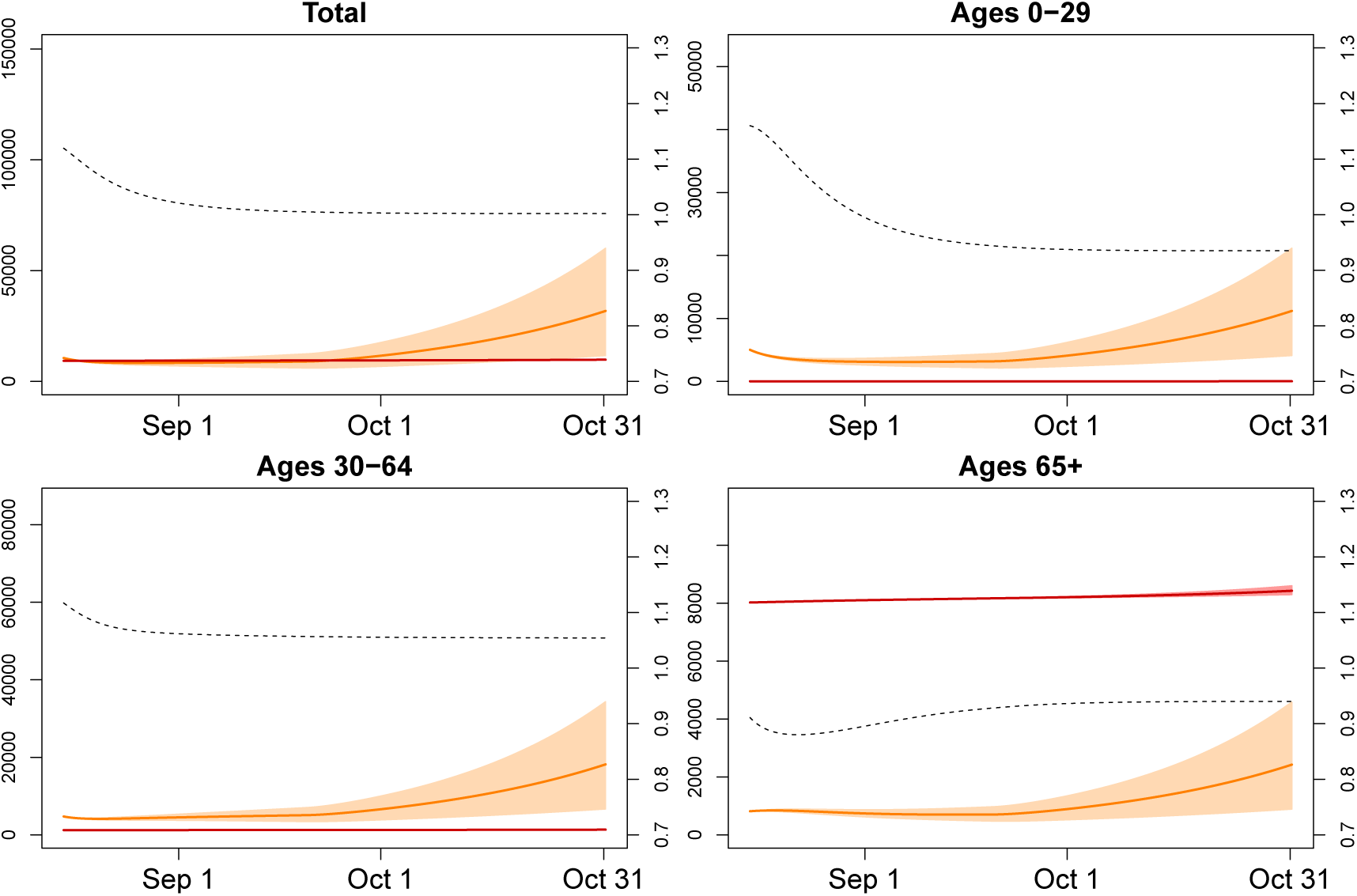
Scenario 2: Infection numbers for different age groups (solid orange line: mean active infections at time t, solid red line: mean cumulative number of deaths until time *t*, intervals represent 80% range due to parameter uncertainty, dashed line: male/female ratio of infections at time *t*).

**Figure 7:**
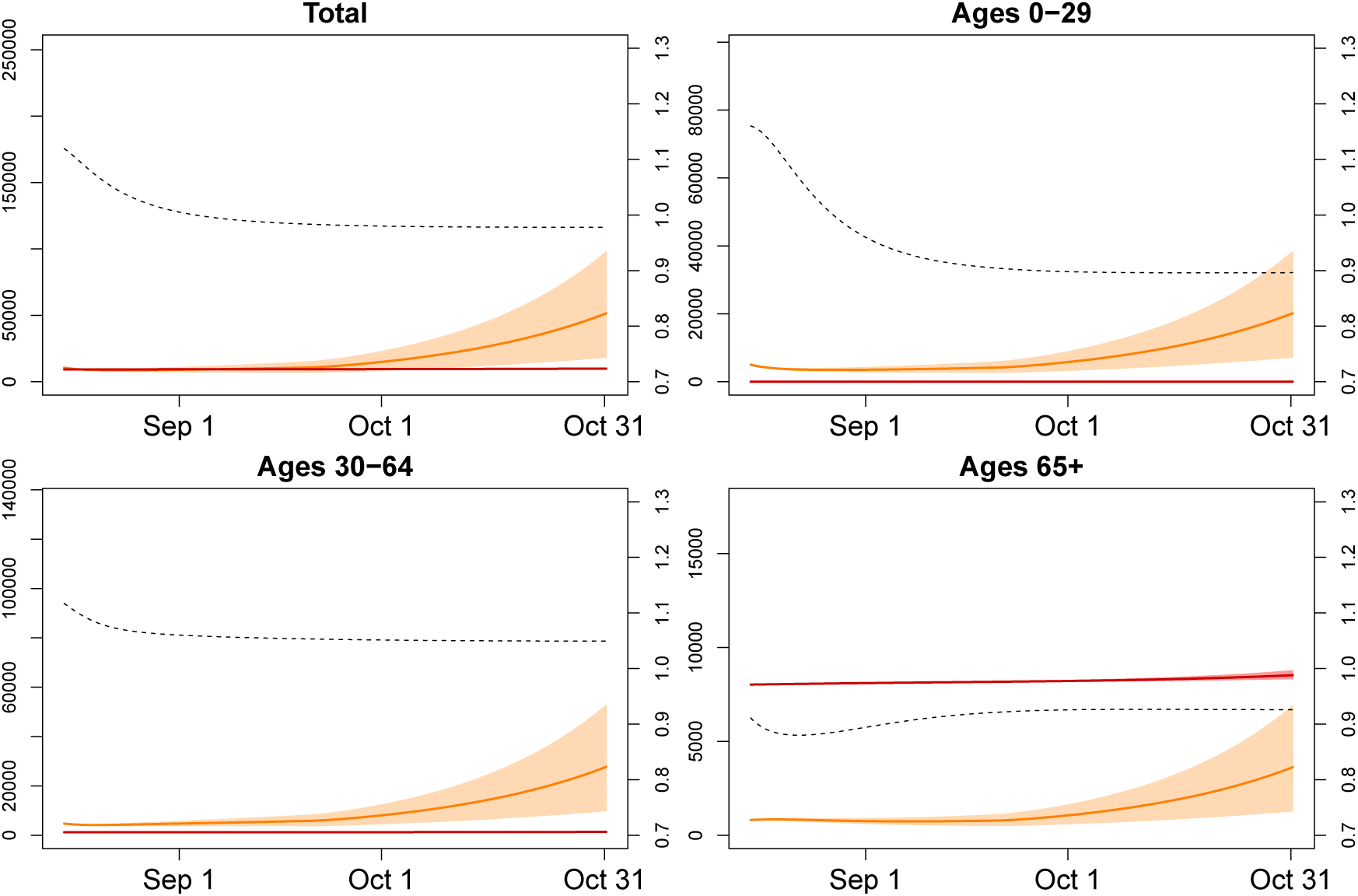
Scenario 3: Infection numbers for different age groups (solid orange line: mean active infections at time t, solid red line: mean cumulative number of deaths until time *t*, intervals represent 80% range due to parameter uncertainty, dashed line: male/female ratio of infections at time *t*).

**Figure 8:**
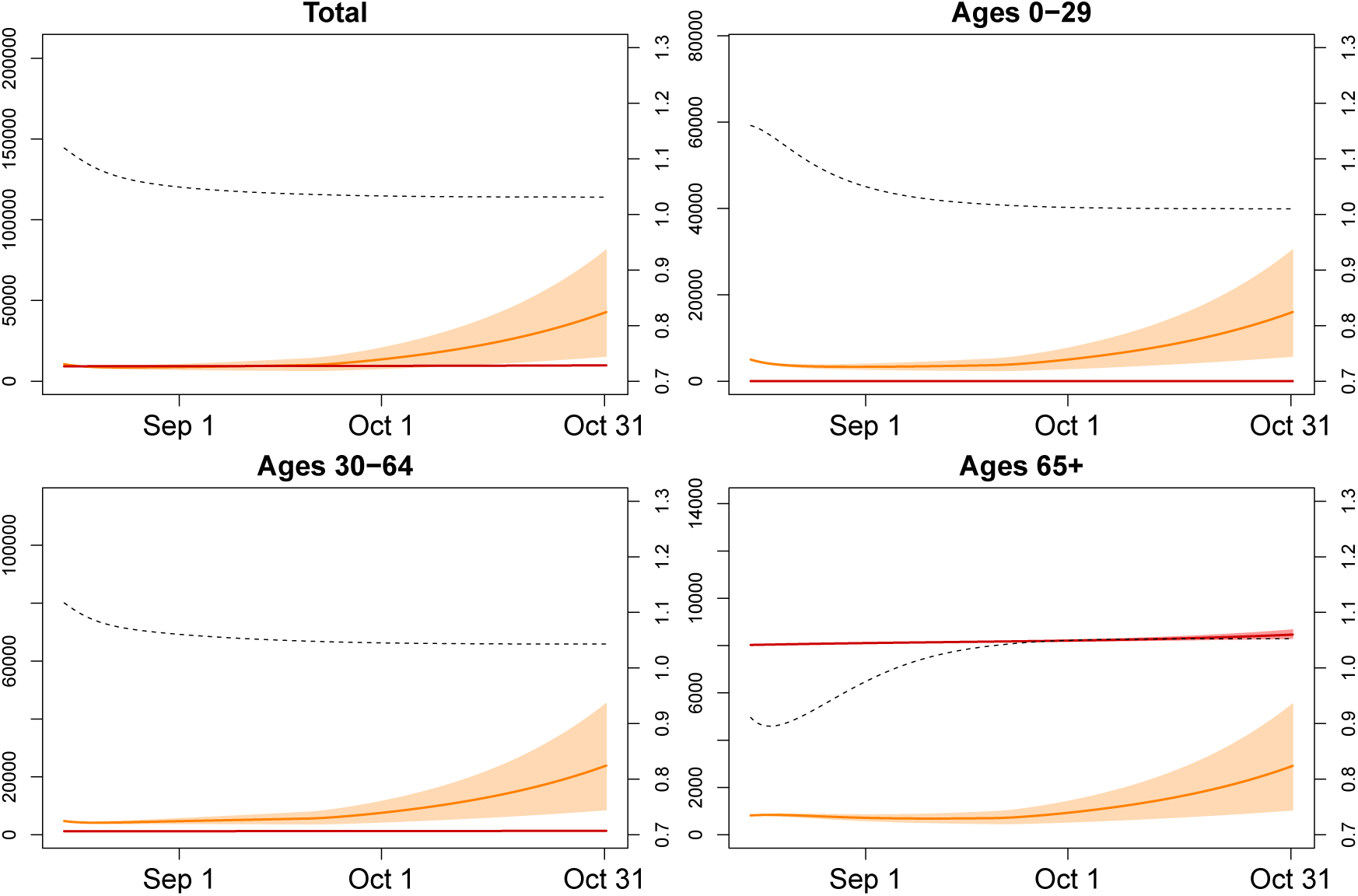
Scenario 4: Infection numbers for different age groups (solid orange line: mean active infections at time t, solid red line: mean cumulative number of deaths until time *t*, intervals represent 80% range due to parameter uncertainty, dashed line: male/female ratio of infections at time *t*).

**Figure 9:**
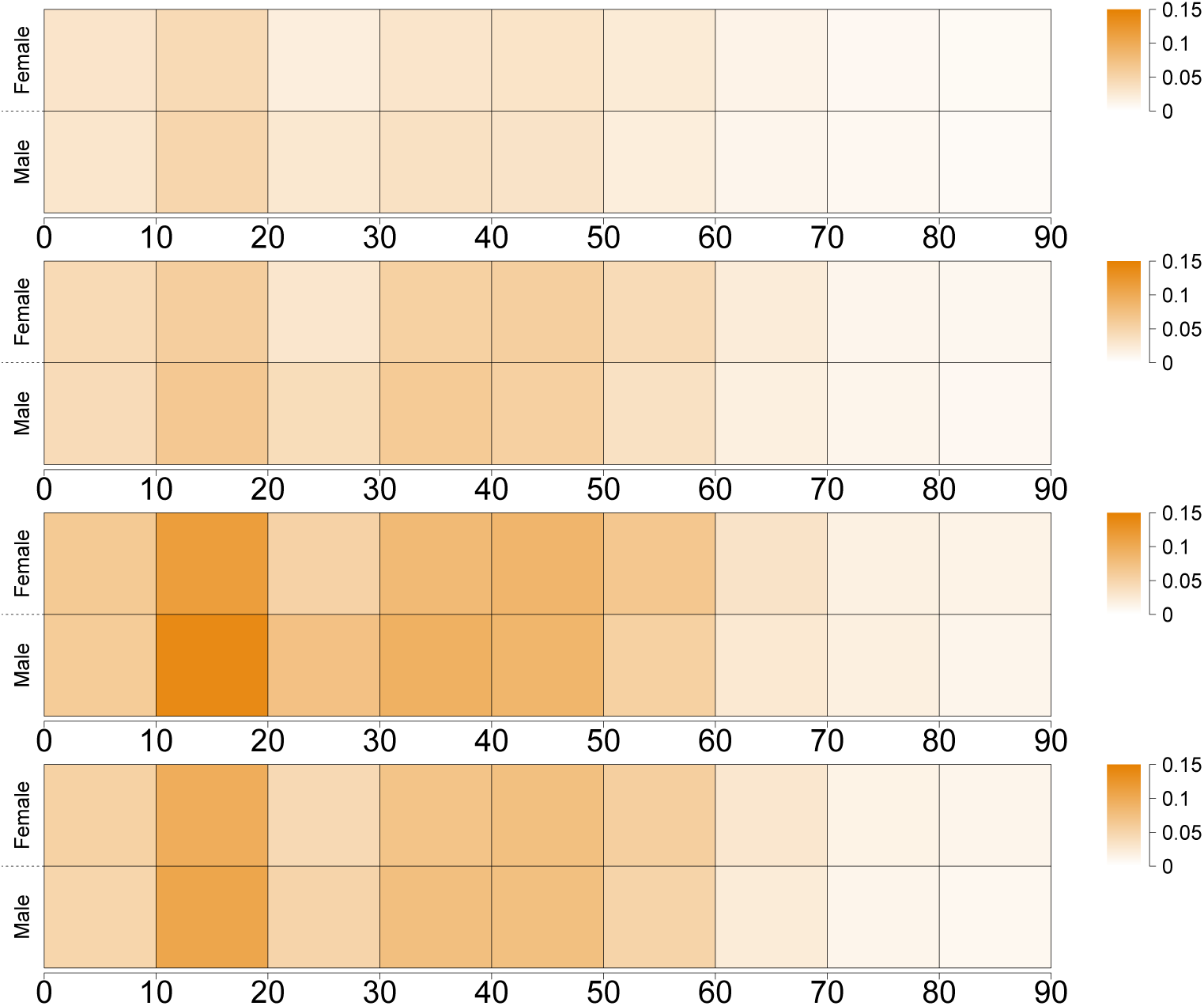
Incidence of active infections for different sex and age groups in scenarios 1–4 (scale is in % per sex and age group).

The behaviour of the epidemiological model is primarily governed by the effective contact rates *β*_*ab*_ which result from the basic contact rates *λ*_*ab*_ by accounting for the secondary attack rate and lockdown measures. It is implicitly assumed here that hospitalised cases are effectively isolated from the remaining population and can not spread the disease. Note that the product (1 − *m*_*ab*_)(1− *r*)(1− *h*_*b*_) represents the proportion of potential virus transmissions that are not prevented.

## 3 Numerical implementation

We have implemented the suggested model in R using a discrete approximation of the ODE system via the Forward Euler Method (see Butcher (2016)). The step size Δ*t* is chosen as a quarter fraction of one day. Accordingly, the transition rates between the compartments need to be adjusted, whereas the fraction parameters remain unchanged. For instance, if the average incubation time is 5 days and Δ*t* = 1*/*4 (days), the transition parameter *ϵ* = 1*/*5 · 1*/*4 = 1*/*20, whereas the manifestation index *ρ*, as the relative proportion of exposed people developing symptoms, is the same for any Δ*t*. The time-discrete approximation of the system of ODEs is therefore described as follows.

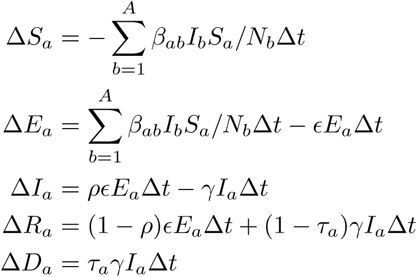

For the involved epidemiological parameters, estimates are available from Imperial College COVID-19 Response Team (2020) and Verity et al. (2020). Pastor-Barriuso et al. (2020) provide estimates of the age- and sex-specific infection fatality rates, based on an seroepidemiological study.

### 3.1 Model fitting

We suggest to fit the model along the following consecutive steps:

1. Determine a timespan {1, …, *T*} during which no lockdown measures had been in place, and determine the cumulative number of infections during this time.
2. Based on plausible ranges for the involved compartment parameters and the initial state of the compartment model, fit the contact intensity model with regard to the cumulative number of infections during {1, …, *T*}.

In order to derive the secondary attack rate *w* from the contact rates *λ*_*ab*_ given in van de Kassteele et al. (2017), we fit the proposed compartment model to the reported cases during a timespan {1, …, *T*} of no lockdown. This step is necessary, because the social contact rates *λ*_*ab*_ do not incorporate the specific transmission characteristics of SARS-CoV-2, such as the average length of the infectious period and average infection probability per contact. We assume that *w* is not specific to age or sex. We employ

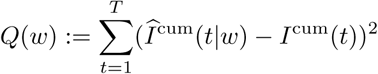

as a least-squares criterion function in order to determine the optimal value 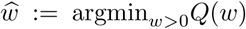,where *I*^cum^ are the observed cumulative infections, and 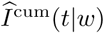 are the estimated cumulative infections based on the epidemiological model given *w*. Hence, 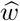 is the scalar parameter for which the cumulative infections are best predicted retrospectively. Note that the observed cumulative number of infections is usually recorded for each day, while the step size Δ*t* in the model may be different. Thus, appropriate matching of observed and estimated values is necessary.

This fitting method requires that the number of infections for the geographical region considered is sufficiently large, such that the mechanics of the compartment model are plausible. Note that potential under-ascertainment may not substantially change the optimal value of *w* as long as the proportion of detected cases does not strongly vary over time. Furthermore, the suggested fitting method is based on the assumption that the probability of virus transmission is independent of age and sex, given that a contact has occurred. If different propensities of virus transmission are allowed for, the contact matrix may be correspondingly adjusted along introduced parameters *w*_1_, …, *w*_*ab*_ for each group combination or *w*_1_, …, *w*_*a*_, if the probability of transmission only depends on the contact group. The criterion function is likewise extended as (*w*_1_, …, *w*_*ab*_) ↦ *Q*(*w*_1_, …, *w*_*ab*_). However, optimisation in this extended model requires a sufficiently large number of transmissions and detailed information on the recorded infections, and may lead to unpractically vague estimates otherwise. Therefore, we suggest to employ the simpler model with univariate *w* first.

### 3.2 Sensitivity analysis and parameter uncertainty

In order to account for parameter uncertainty, we develop uncertainty intervals for the number of people in each compartment. As a cautionary remark, note that these intervals are not to be equated to confidence intervals in the classical sense. Though the resulting intervals are conceptually comparable to Bayesian credibility intervals, they are to be distinguished in that no prior distribution is explicitly assumed here. Note that these intervals do not reflect uncertainty in terms of the underlying infection data.

We predict the number of cases in each age-specific compartment using a Monte Carlo simulation method. For each simulated run, all parameters are independently drawn from their respective range, yielding an instance of a hypothetical parameter setup. Given these parameters, the SEIRD ODE model is approximated using the Forward Euler Method and known initial states, as described above. After *N*_*R*_ of such simulated runs, the prediction intervals for all relevant values are construed based on the pseudo-empirical trajectories of the compartment model. Furthermore, prediction intervals are derived as point-wise quantile ranges for each *t*. For instance, an 80% prediction interval for the number of infectious people in group *a* at time *t* is [*I*_*a*,10%_(*t*), *I*_*a*,90%_(*t*)].

## 4 Analytical approach and scenarios

First, we fitted the model to observed COVID-19 infections using transition rates from literature as described under Section 2 for the period 21 February to 13 March 2020. We estimated the model parameter *w*, also termed secondary attack rate, which reflects the probability of infection per contact, by least squares between observed and predicted values, as described in Section 3.1. Second, we developed four scenarios starting our projections on 15 August 2020 and, using quarter-days as base time, ending on 31 October 2020. The first scenario, which is our baseline scenario, assumed that the age- and sex-specific contacts are down by 80%, i.e. only 20% of the contacts estimated by van de Kassteele et al. (2017) were realized between start and end of the projection. This applied to all age groups and to both sexes. This scenario should reflect continuous distancing measures as were present in mid-August. The second scenario assumed that contacts at working ages 30–59 were increased by 5 percentage points (PP), and among those aged 60–69 by 2.5 PP, equaling a decline of 76% and 78% respectively. All other ages remained at 80% contact reduction. This should reflect the return from home office settings, the opening of shops, cafes, restaurants, etc. The third scenario considers an additional increase in contact rates among ages 10–29 by 5 PP, which should reflect the opening of schools and venues mainly visited by young individuals. Scenario 4 explores the impact of sex-specific contacts by aligning the female contacts to the level of male contacts. We explored the following age-specific outcomes:

1. Number of active infections which were defined as the number of individuals in compartment *I* by age and sex,
2. Cumulative number of deaths out of compartment *I* by age and sex,
3. Excess number of deaths in scenarios 2, 3 and 4 in comparison to scenario 1 by age and sex,
4. Sex ratio of incidence defined as male/female ratio of the number of new COVID-19 cases divided by the total population (*S*_*a*_)
5. Sex ratio of mortality rate defined as male/female ratio of the number of deaths out of compartment *I* divided by the total population (*S*_*a*_).

## 5 Results

Fitting our model to COVID-19 infections observed during our fitting period (21 Feb – 31 March 2020) results in an estimate of the secondary attack rate *w ≈*13%. We started with 10,572 active infections on 15 August 2020 and under Scenario 1 this figure increased to approximately 20,115 (Figure 3) (men: 9,960; women: 10,155). The number of active infections was highest at ages 30–39 (men: 1,705; women: 1,885), followed by ages 10–19 (men: 1,676; women: 1,770), and ages 40–49 (men: 1,628; women: 1,619). The cumulative number of deaths increased from 9,258 to 9,687 with 5,416 men and 4,271 women. By 31 October 2020, infection rates (Table 1) were highest among the 10–19-year old (men 43.4 and women 48.5 per 1000 individuals) followed by ages 30 to 49 (above 30 for both genders), and ages 0–9 (around 30 for both genders). At ages above 50, infection rates declined rapidly, almost halving from individuals in their fifties (men: 24.0; women: 20.4) to those in their sixties (men: 13.5; women: 11.2), while at older ages the decline followed at a much lower pace (ages 70–79: men: 7.7; women: 8.1; ages 80+: men: 6.2; women: 5.3). Sex ratios of infections were below 1 in the age interval 10 to 49, indicating a higher risk of infections among women. From age 50 onwards they were generally above 1 (with the exception of ages 70–79), thus turning the disadvantage towards men. As expected, death rates (Table 1) increased exponentially with age. They were more than twice to three times as high among men than women.

**Table 1:** Mean infection rates, death rates (in %) and male/female ratio. The infection rates are with respect to projected active infections on 31 Oct 2020. The death rates are with respect to the considered timespan 15 Aug 2020 to 31 Oct 2020, and are scaled to annual rates.

| **Mean infection rates on Oct 31 (in %)** |  |  |  |  |  |  |  |  |  |  |
| --- | --- | --- | --- | --- | --- | --- | --- | --- | --- | --- |
|  |  | 0–9 | 10–19 | 20–29 | 30–39 | 40–49 | 50–59 | 60–69 | 70–79 | 80+ |
| Scenario 1 | female | 0.0299 | 0.0485 | 0.0276 | 0.0356 | 0.0324 | 0.0204 | 0.0112 | 0.0081 | 0.0053 |
|  | male | 0.0313 | 0.0434 | 0.0200 | 0.0304 | 0.0321 | 0.0240 | 0.0135 | 0.0077 | 0.0062 |
|  | ratio | 1.05 | 0.89 | 0.73 | 0.86 | 0.99 | 1.18 | 1.21 | 0.94 | 1.17 |
| Scenario 2 | female | 0.0419 | 0.0644 | 0.0405 | 0.0616 | 0.0550 | 0.0354 | 0.0179 | 0.0119 | 0.0077 |
|  | male | 0.0438 | 0.0572 | 0.0294 | 0.0546 | 0.0564 | 0.0424 | 0.0222 | 0.0114 | 0.0092 |
|  | ratio | 1.04 | 0.89 | 0.73 | 0.89 | 1.03 | 1.20 | 1.24 | 0.96 | 1.20 |
| Scenario 3 | female | 0.0604 | 0.1379 | 0.0726 | 0.0922 | 0.0864 | 0.0544 | 0.0262 | 0.0180 | 0.0120 |
|  | male | 0.0626 | 0.1147 | 0.0526 | 0.0813 | 0.0868 | 0.0657 | 0.0324 | 0.0170 | 0.0141 |
|  | ratio | 1.04 | 0.83 | 0.73 | 0.88 | 1.00 | 1.21 | 1.24 | 0.95 | 1.17 |
| Scenario 4 | female | 0.0487 | 0.1052 | 0.0501 | 0.0753 | 0.0747 | 0.0505 | 0.0229 | 0.0116 | 0.0088 |
|  | male | 0.0535 | 0.0967 | 0.0442 | 0.0697 | 0.0746 | 0.0565 | 0.0280 | 0.0145 | 0.0120 |
|  | ratio | 1.10 | 0.92 | 0.88 | 0.93 | 1.00 | 1.12 | 1.22 | 1.24 | 1.37 |

| **Mean death rates until Oct 31 (in %)** |  |  |  |  |  |  |  |  |  |  |
| --- | --- | --- | --- | --- | --- | --- | --- | --- | --- | --- |
|  |  | 0–9 | 10–19 | 20–29 | 30–39 | 40–49 | 50–59 | 60–69 | 70–79 | 80+ |
| Scenario 1 | female | 0.0000 | 0.000 | 0.0001 | 0.0003 | 0.0003 | 0.0009 | 0.0015 | 0.0048 | 0.0086 |
|  | male | 0.0000 | 0.000 | 0.0000 | 0.0001 | 0.0006 | 0.0020 | 0.0049 | 0.0111 | 0.0246 |
|  | ratio | – | – | 0.4000 | 0.3800 | 1.8900 | 2.2300 | 3.2600 | 2.3200 | 2.8500 |
| Scenario 2 | female | 0.0000 | 0.000 | 0.0001 | 0.0004 | 0.0005 | 0.0013 | 0.0020 | 0.0058 | 0.0100 |
|  | male | 0.0000 | 0.000 | 0.0000 | 0.0002 | 0.0009 | 0.0029 | 0.0065 | 0.0134 | 0.0293 |
|  | ratio | – | – | 0.3900 | 0.3900 | 1.9400 | 2.2700 | 3.3300 | 2.3400 | 2.9200 |
| Scenario 3 | female | 0.0000 | 0.000 | 0.0001 | 0.0005 | 0.0006 | 0.0016 | 0.0024 | 0.0072 | 0.0125 |
|  | male | 0.0000 | 0.000 | 0.0000 | 0.0002 | 0.0012 | 0.0037 | 0.0081 | 0.0165 | 0.0360 |
|  | ratio | – | – | 0.3800 | 0.3800 | 1.9100 | 2.2900 | 3.3400 | 2.3000 | 2.8900 |
| Scenario 4 | female | 0.0000 | 0.000 | 0.0001 | 0.0004 | 0.0006 | 0.0016 | 0.0023 | 0.0052 | 0.0102 |
|  | male | 0.0000 | 0.000 | 0.0000 | 0.0002 | 0.0011 | 0.0034 | 0.0074 | 0.0151 | 0.0331 |
|  | ratio | – | – | 0.4600 | 0.4000 | 1.9000 | 2.1400 | 3.2900 | 2.9200 | 3.2500 |

**Table 2:**
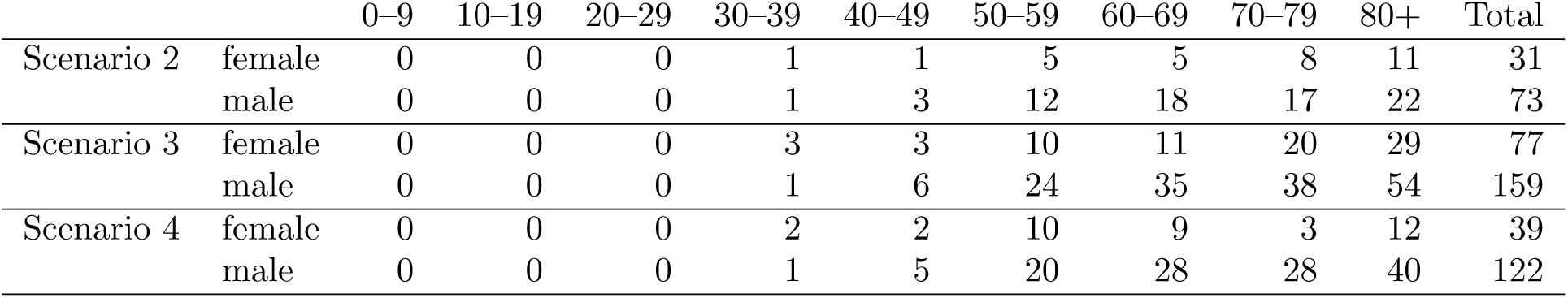
Mean excess number of deaths in Scenarios 2–4 in comparison to Scenario 1 until 31 Oct 2020.

Scenario 2 assumed increased contacts at working ages and arrived at 31,800 active infections by 31 October 2020 and therefore 11,685 active infections more than in scenario 1 (men 5,730; women 5,955). These additional infections stemmed from all ages, even if the risk of infections increased most among the working ages. Sex ratios of infection rates turned toward the disadvantage of men from age 30 onwards. The additional infections translated into an additional 104 deaths (men: 73; women: 31); among women, 61% of these deaths resulted at ages 70 and above; among men, 53%, reflecting their higher mortality already at younger ages. Also the sex ratios of death rates turned towards the disadvantage of men. Scenario 3 with increased contacts at young and working ages resulted in 51,521 active infections and thus 31,406 more than in Scenario 1 (men: 15,894; women: 15,512) which translated into an additional 236 deaths with the majority resulting from ages 70 and above (men: 57%; women: 63%). Sex ratios, both in infections and deaths, became even more unfavourable for men.

Scenario 4 used similar assumptions as scenario 3 but the contact rates of women were lowered to those of men. This translated into 42,838 active infections which are 22,723 more than in scenario 1, but 8,683 less than in scenario 3. More infections were spared among men (−4,960) than among women (−3,725). While the number of excess deaths was still higher than in scenario 1 (men: 122; women 39), it was lower than in scenario 3 (men: 122 −159 = −37; women: 39− 77 = −38). Thus, in absolute terms men profited as much as women.

## 6 Discussion

Incorporating age- and sex-specific contact rates in a COVID-19 compartment model permits exploration of the effects of changes in mitigation measures on the two genders. We developed four scenarios which assumed ongoing distancing measures versus easing of contact restrictions in working ages, and among adolescents and young adults. Our projections do not set out to forecast the actual number of COVID-19 infections in a time span of about two months, they rather assess the effect of increased contacts on the infection and mortality risks of the two genders and the various age groups. The fit of our model to the baseline period in February and March results in an estimated secondary attack rate *w ≈*13%, putting our findings in close agreement with the rates reported in Ghangdou, where the household *w* varied between 12% and 17%, and the non-household *w* between 6% and 9% (Jing et al. (2020)), although higher attack rates of up to 35% have been reported e.g. for meals and holiday visits (Liu et al. (2020b)). Three important lessons can be learned from our scenarios.

First, even a small change in contact rates has a large impact on infections and deaths. In our projections we assumed an increase ranging from 5 to 2.5 PP. This reflects the fact that without non-pharmaceutical mitigation measures (NPMM) such as masks, physical distance between individuals, better air ventilation and hygiene, and without contact tracing, the infection rates would return to the initial exponential increase. This was reflected in a reproduction rate of 3.3 to 3.8, as observed at the beginning of the pandemic (Lin et al. (2020), Liu et al. (2020) and Alimohamadi et al. (2020), RKI (2020)). However, the presence of NPMM also mitigates the effect of the increase in contacts due to the return to office, opening of shops, restaurants, as well as schools, and venues visited by young adults, leaving it far from the initial impact. In our present scenarios, both effects, the change of contact rates and the change of their impact, are captured in the reduction matrix (*m*_*ab*_), which is multiplied with the matrix of the contact rates. One alternative approach would be to develop separate scenarios for changes in the secondary attack rate *w* due to NPMM and changes in the contact rates (*m*_*ab*_), which is one possibility to modify this analysis further. At any rate, our scenarios show that small changes already have large impacts on infections and deaths. This implies that the impact of contacts must be diminished considerably to allow increases in contacts without returning to exponential growth of infections, hence underlining the high importance of the NPMM in the current phase of the pandemic.

Second, due to intergenerational contacts, any easing of measures in working and young ages will inevitably lead to an increase in infections and deaths at all ages. People at old ages will suffer most with elderly men being at a particular high risk of death due to increased contacts. Most interestingly, this increased mortality is also transmitted by the higher contact rates of women, as shown in our scenario 4. Mortality may have changed over the course of the pandemic because of better treatment options of critically severe COVID-19 cases using, e.g., dexamethasone (Cain and Cidlowski (2020)). Our mortality rates based on Pastor-Barriuso et al. (2020) are based on Spanish data from 27 April – 22 June 2020, which already should reflect a possible decline. Our results emphasise that increases in contacts need to be accompanied by special measures protecting the elderly from death, without negative physical and mental health consequences due to quarantine and isolation measures (Galea et al. (2020)). Contrary to deaths, infections will mainly increase at young and middle ages with a lower risk of severe COVID-19 symptoms or even asymptotic disease courses.

Third, small increases in contact rates change the sex ratios in infections and deaths towards the disadvantage of men. At all ages, men will have more than twice the mortality risk from COVID-19, while the risk of infections is more frequent among working age women than men. At old ages, men have higher infection risk. Note that, in absolute numbers, more women are diagnosed with COVID-19 at old age due to their higher life expectancy. Here a more substantial question arises, namely whether COVID-19 infection rates are indeed gender-specific. German COVID-19 infection rates, as in any other country, are biased by the time-lag of reporting and by differential availability of PCR tests over time and to subgroups of the population (RKI (2020)). Gender-specific diagnoses in favour of women may reflect that higher contact intensities of women may have led to a higher rate of conducted PCR tests and therefore to a smaller number of undiagnosed cases. In addition, women are more health-conscious than men (Oksuzyan et al. (2020)), may have sought PCR testing to a higher degree even when presented with weaker symptoms, and are more adherent to NPMM (Galasso et al. (2020)). On the other hand, Takahashi et al. (2020) found sex-specific differences in immune response to COVID-19 infections. For a further discussion of potential sex-specific mechanisms modulating the course of disease, see also (Gebhard et al. (2020)). Thus, we can conclude that both biological and social factors contribute to sex- and gender-specific infection and mortality rates.

A sizeable proportion of infections and deaths is transmitted through the higher contact rates of women, as shown in our scenario 4. This higher number of contacts may primarily result from care obligations where women are the main care providers. By mid-July, among the COVID-19 infection cases reportedly cared for or working in medical facilities, 72% were women and 28% men with a median age of 41 years (RKI (2020)). Since women have a higher untapped work-from-home capacity than men (Alipour et al. (2020)) better exploitation of their work-from-home potential may safe infections and lives.

We focused on the practical emulation of the dynamic behaviour and process of the spreading of COVID-19 while incorporating specific epidemiological information on the virus and disease. To achieve this aim we used a compartment modeling framework, which has become a standard approach in epidemiology due to its flexibility and accessibility. The main advantage of this modeling framework is that a considerable amount of demographic and epidemiological information can be incorporated while the essential model structure and implementation remain relatively simple. Similarly, it is possible to extend the model to incorporate parameter uncertainty, as described above. Furthermore, we want to emphasize the Markov-like property of compartment modeling in the sense that current compartment sizes on a specific date are sufficient for deducing the subsequent behaviour of the epidemiological process, which makes the framework particularly attractive for forecasting and investigating hypothetical scenarios. However, there is one drawback to compartment modelling that it is inherently based on an averaging rationale which treats population groups homogenously and the average number of contacts in each group is a determining parameter. In contrast to truly stochastic models (such as agent-based models), no random or systematic individual deviations from the fundamental contact patterns are taken into consideration. Likewise, compartment modeling is not suitable for assessing local dynamic behaviour, such as the notions of infection clusters and superspreading events. In addition, geographical and spatial information are not explicitly considered in compartment modeling, and this further limits the scope of the forecasting results. In general, assessing the impact of introducing or easing different lockdown measures is remarkably difficult, especially because several aspects are usually changed simultaneously and the general behaviour of the population may change dynamically at the same time. Some efforts have been made to address these issues in the literature, however we advise against using the proposed model for such purposes. One main reason is that the initial state for forecasting and fitting of the model relies primarily on available data sources, which are in the form of reported count data. In addition to the general limited validity of observational data, there is still insufficient knowledge on the specific characteristics of COVID-19 and the actual current spread of the virus. Naturally, other modeling approaches face the same issues of data quality.

In our COVID-19 forecasts, the number of infections and the number of deaths differ only slightly from models which do not differentiate by sex (data not shown). However, age- and sex-specific models provide better insight into the risk populations of infections and mortality. This helps to target health policy measures under scarce resources, such as who should be tested and vaccinated first. Both biological sex and social gender appear to affect COVID-19 infection rates and their outcomes; this needs to be acknowledged in health policy decisions and medical treatment. To further explore social factors on COVID-19 transmission, more information that includes socio-demographic data is needed.

## Data Availability

All data that has been used in the manuscript is publicly available.
An R implementation of the proposed model is available via Github:
https://github.com/AchimDoerre/Covid-19

## Notes

### Competing Interest Statement

The authors have declared no competing interest.

### Funding Statement

No external funding, payment or services have been received for any aspect of the submitted work.

### Author Declarations

The article concerns numerical studies and no trials/experiments involving humans have been conducted.

### Summary of Updates

Section 3 on numerical aspects has been shortened to clarify main numerical issues. In Section 4 and 5, we add an additional scenario. Minor numerical inaccuracies have been corrected in the implementation. All figures in the results section 5 have been revised.

